# Associations Between Measures of Sleep Health and Cognitive Function in Older Adults

**DOI:** 10.64898/2026.09.21.26363558

**Authors:** Kristen L. Knutson, Katharine Harrington, Shaina J Alexandria, Phyllis C Zee, Mercedes R. Carnethon

## Abstract

**Objectives:** Prior research has identified associations between sleep health and cognitive function. Our aim was to investigate multiple domains of sleep health and their associations with cognitive function in a diverse sample of older adults.

**Methods:** These data are from an observational study in Chicago, USA, that enrolled adults aged 55 years and older without cognitive impairment at baseline and followed a subset for up to 2 years. Sleep was measured with 7 days of wrist actigraphy and we examined sleep duration (amount), sleep fragmentation (an index of restlessness and sleep quality), sleep start time and sleep regularity (based on the within-subject standard deviation of sleep start time)..

Participants also completed tasks from the NIH Toolbox to estimate fluid cognition and the Trail Making Test (Trails B) to assess attention and processing speed. A subset of participants was examined at a follow-up visit that included only the NIH Toolbox assessment.

**Results:** Our sample included 324 participants (63.6% women, 37.3% African American/Black, 13.6% Hispanic/Latino). Greater sleep fragmentation and less sleep regularity were associated with worse fluid cognition and Trails B scores (all p<.01). Longer sleep duration and earlier sleep start times were associated with better performance on Trails B. No sleep measures were associated with change in the fluid cognition score over an average of 1.9 years (n=133).

**Discussion:** These findings support a cross-sectional association between cognitive function and multiple domains of sleep health, including duration, quality, timing and regularity, in a diverse sample of older adults.

## Introduction

Cognitive health is a critical component of healthy aging and quality of life for older adults, while significant cognitive decline can lead to loss of independence and increased disability. Therefore, identification of modifiable behaviors or conditions associated with cognitive function in older adults can inform strategies to maintain cognitive health and slow cognitive decline with aging. Sleep is one modifiable lifestyle behavior that has been linked to cognitive health.

Healthy sleep comprises several domains, including adequate duration or amount of sleep, good quality sleep, optimal timing of sleep, and regular timing from day to day. Prior studies have found associations between characteristics of healthy sleep and cognitive function. For example, a study of women 65 years and older measured sleep objectively using wrist actigraphy and found that poor sleep quality was associated with worse cognitive function in both crosssectional [1] and longitudinal [2] analyses. A separate study conducted among older men (≥65 years) also found that worse sleep quality assessed via wrist actigraphy was associated with worse cognitive function in cross-sectional and prospective analyses [3, 4]. A large study among Hispanic/Latino adults aged 45-65 years found that greater sleep regularity, but not sleep quality nor timing, based on wrist actigraphy was associated with worse cognitive function [5]. Thus, these studies, which all originate from specific demographic groups (men, women or Hispanic/Latino adults) identified some domains of sleep health that were associated with cognitive function.

The goal of our study was to examine multiple dimensions of sleep health, measured objectively via wrist actigraphy, in a racially diverse sample of adults 55 years and older and determine which of these dimensions were associated with cognitive function in both cross-sectional and longitudinal analyses.

## Methods

### Sample

These data are from the Disparities in Sleep and Cognitive Outcomes (DISCO) study, an observational cohort study of adults aged 55 years or older [6]. Inclusion criteria included age of 55 years or older, self-reported race/ethnicity of black, white or Hispanic/Latino/a/x; ability to speak English or Spanish, community dwelling (e.g. not in assisted living or health facility), and a Montreal Cognitive Assessment score (MoCA) ≥23 in-person or blind MoCA≥18 (over the phone) to rule out cognitive impairment at baseline. This observational study included objective estimates of sleep via 7 days of wrist actigraphy as well as cognitive testing that assessed multiple domains of cognitive function. The study included a baseline assessment (Visit 1) as well as a longitudinal follow-up (Visit 2) in a subset of participants. The protocol was reviewed and approved by the Institutional Review Board at Northwestern University and all participants gave written informed consent.

### Measures

#### Sleep measures

The primary measures of sleep health were obtained using a wrist actigraphy device over 7 days (Actiwatch Spectrum Plus, Phillips). These included participants’ average ***sleep duration*** (amount of sleep obtained per night), average ***sleep fragmentation*** (an index of restlessness and sleep quality), average ***sleep start time*** (indicator of sleep timing), and ***start time standard deviation*** (lower values indicate greater sleep regularity).

#### Cognitive tests

Cognitive outcomes were obtained from the NIH Toolbox and Trail Making Test Part B (Trails B) cognitive exams at Visit 1 and the NIH Toolbox at Visit 2. The Trails B test was not repeated at Visit 2. The NIH Toolbox assessments included the list sorting task (working memory), Pattern Comparison (processing speed), picture processing (episodic memory), and the Flanker Test (inhibitory control). We calculated a fluid cognition score by averaging the scores of these four NIH Toolbox items (higher values indicate better cognitive function) [7]. The Trails B test involves connecting letters and numbers in sequential order but alternating between letters and numbers. This test assesses attention and processing speed, and higher scores indicate greater time to completion or worse cognitive function. For the secondary, longitudinal analysis, we calculated the change in fluid cognition score from Visit 1 to Visit 2 divided by the time between the two visits.

#### Covariates

Age, race (Black vs. White), ethnicity (Hispanic vs. Non-Hispanic), and sex (Female vs. Male) were included in analyses.

### Statistical analysis

Participants were included if they had at least 4 valid days of actigraphy data, complete covariate data, and at least one cognitive outcome variable (n=324). All sleep measures were centered and scaled to allow interpretation of model estimates as a 1-standard deviation increase in the sleep variable. Participants with extreme sleep start and end times (falling asleep after 2AM or waking before 4AM) were removed in sensitivity analyses (sensitivity sample n=304). A secondary analysis investigated the change in fluid cognition from Visit 1 to Visit 2. Participants from the primary sample who completed Visit 2 were included in the secondary, longitudinal analyses (n=133).

Descriptive statistics for all variables were calculated. Associations between sleep measures and NIH Toolbox fluid cognition score were assessed using multivariable linear regression models. Associations between sleep measures and Trails B scores were assessed using Poisson regression models to account for the skew of the Trails B data. The primary model adjusted for age, race, ethnicity, and sex and in sensitivity analyses, and in sensitivity analyses, we reran these models removing extreme values for sleep start times. The longitudinal analyses also adjusted for age, race, ethnicity, and sex as well as baseline fluid cognition score. Data management and analyses were completed using SAS 9.4 and R.

## Results

Table 1 describes our sample, which included 206 women (63.6%) and 121 (37.3%) African American/Black adults in the baseline visit. Average sleep duration was 409±64 minutes (or 6.8±1.1 hours) and average sleep fragmentation was 22±8.4%. Participants fell asleep at approximately 23:18±1.4 hours, on average. The average fluid cognition score was 92.1±8.7 while the average Trails B score was 90.5±51.2 seconds. For the subset who attended Visit 2, the average fluid cognition score was 92.6±7.5 at Visit 2 and the average change in fluid cognition scores was -0.06 ± 3.4. The average interval between Visit 1 and Visit 2 was 1.80±0.8 years.

**Table 1.** Sample Description.

| **Demographics (n=324)** | **Mean (SD) or N (%)** |
| --- | --- |
| Age (years) | 68.7 (6.25) |
| Gender |  |
| Men, n (%) | 118 (36.4%) |
| Women, n (%) | 206 (63.6%) |
| Race |  |
| African American/Black, n (%) | 121 (37.3%) |
| White, n (%) | 203 (62.7%) |
| Hispanic/Latino, n (%) | 44 (13.6%) |
| **Sleep – Baseline (n=324)** |  |
| Sleep duration (min) | 409 (63.9) |
| Sleep Fragmentation (%) | 21.6 (8.38) |
| Sleep Start Time (h) | 23.3 (1.36) |
| Sleep Start Time SD (min) | 59.0 (34.1) |
| **Cognitive Function – Baseline (n=324)** |  |
| List Sort - Working Memory | 96.7 (11.5) |
| Pattern Comparison - Processing | 87.3 (15.8) |
| Picture Sequence - Memory | 93.5 (12.4) |
| Inhibition Control - Attention | 91.1 (8.72) |
| Fluid Cognition Score | 92.1 (8.73) |
| Trails B (sec) | 90.5 (51.2) |
| **Cognitive Function – Follow-up (n=133)** |  |
| List Sort - Working Memory | 97.1 (10.4) |
| Pattern Comparison - Processing | 88.2 (15.6) |
| Picture Sequence - Memory | 93.6 (10.9) |
| Inhibition Control - Attention | 91.9 (8.22) |
| Fluid Cognition Score | 92.6 (7.54) |
| Change in Fluid Cognition Score | -0.06 (3.35) |

Table 2 presents the results from the regression analyses examining the associations between the measures of sleep health and the cognitive function measures. At baseline, less sleep fragmentation and more regular sleep were both associated with better fluid cognition scores.

**Table 2.**
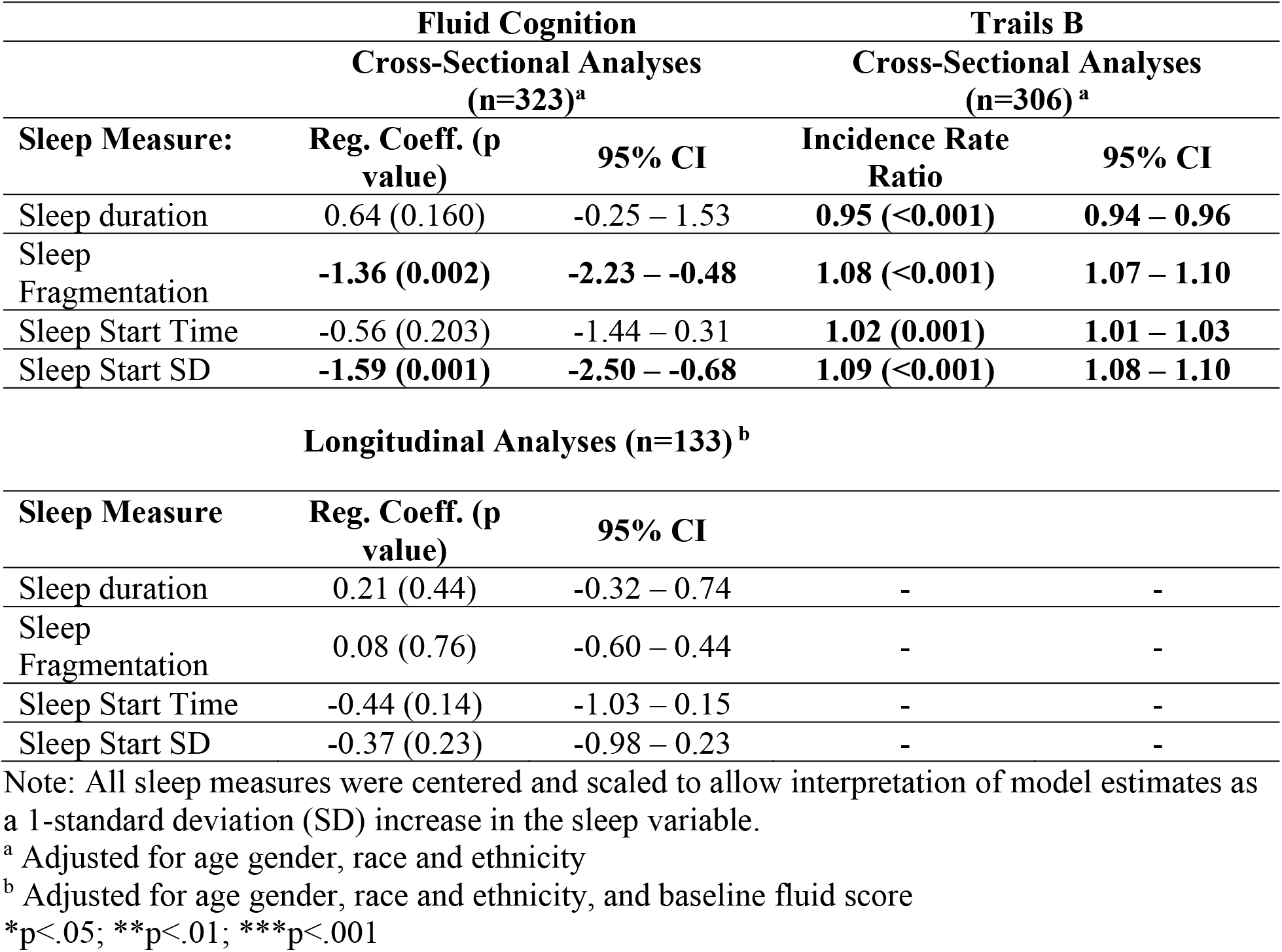
Associations between sleep measures and cognitive function tests.

Specifically, one standard deviation more sleep fragmentation was associated with 1.36 fewer points on the fluid cognition score, while one standard deviation larger sleep variability was associated with 1.59 fewer points on the fluid cognition score. Neither sleep duration nor sleep start time were associated with fluid cognition scores. Better performance on the Trails B task was associated with longer sleep duration, less sleep fragmentation, earlier sleep start time and more regular sleep. Specifically, one standard deviation more sleep duration was associated with 5% faster performance, one standard deviation more sleep fragmentation was associated with 8% slower performance, one standard deviation later sleep start time was associated with 2% slower performance, and one standard deviation more variability was associated with 9% slower performance on Trails B. Analyses that exclude participants with extreme sleep start and end times produced similar associations (results not shown). In the longitudinal subset (n=133), change in the fluid cognition score was not associated with any of the baseline sleep health measures.

## Discussion

In our racially diverse sample of older adults, we found that better fluid cognitive function was associated with better sleep quality and more regular sleep in cross-sectional analyses. We also found that better performance on the Trails B test, which assesses executive function, was associated with all four sleep health domains, including longer sleep duration, better sleep quality, earlier sleep timing and more regular sleep. We did not find any associations between the sleep health measures at baseline and change in fluid cognition scores, however.

Prior observational studies have also demonstrated associations with measures of sleep health based on actigraphy and measures of cognitive function. The Study of Osteoporotic Fractures (SOF) measured sleep via actigraphy in 2932 women aged 65 years and older and measured cognitive function using the Mini-Mental State Examination (MMSE) and the Trails B tests [1]. They found that better sleep quality, based on both greater sleep efficiency and less wake after sleep onset, were associated with better cognitive performance on both the MMSE and the Trails B tests [1]. Greater total sleep time was also associated with better MMSE scores, but not with Trails B [1]. Another large cohort study, MrOS, was comprised of only older men (n=3,132), and also measured sleep via actigraphy and measured cognitive function with a Modified Mini-Mental State examination and the Trails B test [3]. In the MrOS study, less wake after sleep onset was associated with better cognitive performance on the modified MMSE and the Trails B test while greater sleep efficiency was associated with better performance on the Trails B test only [3]. Neither of these studies reported associations with sleep timing or regularity. The Hispanic Community Health Study/Study of Latinos (HCHS/SOL) of 1006 adults aged 45-65 years found that greater sleep regularity was associated with a lower odds of having poor cognitive performance, defined as being >1 standard deviation below the mean on a composite cognitive performance variable based on four tasks [5]. The HCHS/SOL study also examined several measures of sleep timing but none was associated with poor cognitive performance [5]. The cross-sectional associations observed in these studies are consistent with our findings that better sleep quality and greater regularity are associated with better cognitive performance in older adults. Both SOF and MrOS also performed prospective analyses and both observed that worse sleep quality was associated with greater cognitive decline [2, 4], while we did not observe any associations in our prospective analysis. The follow-up period for SOF and MrOS averaged 4.9 and 3.4 years, respectively, both of which were longer than the average follow-up time in our study, which was 1.8 years. It is also important to note that the participants in both SOF and MrOS were predominantly white (≥90% of the sample), while our study has greater racial diversity. Future work should explore whether associations between sleep and cognitive function are modified by sociodemographic factors, such as race, ethnicity and socioeconomic status.

Although these observational studies cannot detect a causal association between sleep and cognitive function, prior research provide biological plausibility for such an association. For example, experimental studies suggest that critical brain maintenance occurs during sleep, such waste clearance [8, 9], or synaptic homeostasis [10]. Thus poor sleep health and sleep disturbances could impair this process and the accumulation of waste products in the brain could impair cognitive function. Another potential mechanistic link between poor sleep and impaired cognitive function involves inflammation. Sleep disturbances are associated with increases in markers of systemic inflammation and neuroinflammation [11, 12], which can lead to cognitive impairment. Sleep disturbances are also associated with changes in autonomic nervous system activity such that sleep loss and sleep disturbances are associated with increased sympathetic relative to parasympathetic activity [13, 14]. The autonomic nervous system helps to regulate cognitive function [15], and greater parasympathetic activity during wake and during sleep is associated with better memory, attention, and executive function [16-20]. Thus, there are several potential pathways through which sleep disturbances could be linked to poorer cognitive function.

There are several strengths to our study, including racial diversity, the objective estimate of multiple domains of sleep health, and assessment of cognitive function using multiple instruments. However, there are some important limitations to note. Our prospective analysis had a smaller sample size, which limits statistical power, and the average follow-up time was just under 2 years. A longer follow-up period may be necessary to detect sufficient change in cognitive function and indeed the average change in fluid cognition in our sample was quite small (-0.06 points). We also did not have repeated Trails B tests at follow-up and were unable to assess prospective associations for this measure, which demonstrated more significant associations with sleep health measures in the baseline cross-sectional analyses. Finally, a causal relationship between sleep health and cognitive function cannot be inferred from these analyses.

The findings from our study provide additional support for an association between multiple domains of sleep health, including duration, quality, timing and regularity, and cognitive function in a diverse sample of older adults. Future research should involve longer prospective studies as well as intervention studies that determine whether improving sleep health, including multiple components of sleep health, can help to maintain cognitive health as we age.

## Data Availability

All data produced in the present study are available upon reasonable request to the authors.

## Notes

**Funding:** This work was supported by National Institute of Aging at the National Institutes of Health (1R01AG059291-A1).

**Data Availability:** The data and materials that support the findings of this study are available from the corresponding author on request. The study reported in this article was not preregistered.

**Conflict of Interests:** Dr. Phyllis Zee is a consultant for Eisai, Takeda, Alkermes, and Jazz and is a Board member of the American Brain Foundation. None of these groups was involved in this study.

### Competing Interest Statement

The authors have declared no competing interest.

### Author Declarations

IRB of Northwestern University gave ethical approval for this work.

